# Pathogenic variants in *SMARCA5*, a chromatin remodeler, cause a range of syndromic neurodevelopmental features

**DOI:** 10.1101/2020.10.26.20217109

**Authors:** Dong Li, Qin Wang, Naihua N. Gong, Alina Kurolap, Hagit Baris Feldman, Nikolas Boy, Melanie Brugger, Katheryn Grand, Kirsty McWalter, Maria J. Guillen Sacoto, Emma Wakeling, Jane Hurst, Michael E. March, Elizabeth J. Bhoj, Małgorzata J.M. Nowaczyk, Claudia Gonzaga-Jauregui, Mariam Mathew, Ashita Dava-Wala, Amy Siemon, Dennis Bartholomew, Yue Huang, Hane Lee, Julian A Martinez, Eva M.C. Schwaibold, Theresa Brunet, Daniela Choukair, Lynn S. Pais, Susan M White, John Christodoulou, Dana Brown, Kristin Lindstrom, Theresa Grebe, Dov Tiosano, Matthew S. Kayser, Tiong Yang Tan, Matthew A. Deardorff, Yuanquan Song, Hakon Hakonarson

**Author notes:** These authors contributed equally to this work (Dong Li, Qin Wang). Correspondence (D.L.), (Y.S.).

## Abstract

Intellectual disability (ID) encompasses a wide spectrum of neurodevelopmental disorders, with many linked genetic loci. However, the underlying molecular mechanism for over 50% of the patients remains elusive. We describe mutations in *SMARCA5*, encoding the ATPase motor of the ISWI chromatin remodeler, as a cause of a novel neurodevelopmental disorder, identifying twelve individuals with *de novo* or dominantly segregating rare heterozygous variants. Accompanying phenotypes include mild developmental delay, frequent postnatal short stature, and microcephaly, and recurrent dysmorphic features. Loss of function of the SMARCA5 *Drosophila* ortholog *Iswi* led to smaller body size, reduced dendrite complexity, and tiling defects in larvae. In adult flies, Iswi neural knockdown caused decreased brain size, aberrant mushroom body morphology and abnormal locomotor function. *Iswi* loss of function was rescued by wild-type but not mutant SMARCA5. Our results demonstrate that *SMARCA5* pathogenic variants cause a neurodevelopmental syndrome with mild facial dysmorphia.

## Introduction

Regulation of gene expression by transcription, requires a highly coordinated and precise process that involves chromatin regulators and modifiers, transcription factors, cohesins, and mediators. The basic and dynamic unit of chromatin is the nucleosome, onto which DNA is wrapped around an octameric histone complex. Subsequent high-order DNA structure requires chromatin remodelers to make DNA accessible, to alter nucleosome position, and in turn, to regulate gene transcription using the energy released by ATP hydrolysis. ATP-dependent nucleosome remodeling not only provides an open and accessible chromatin state, but is also involved in chromatin assembly, nucleosome sliding and spacing, and gene repression, which are crucial for a wide range of biological processes in normal development including early embryonic development stages (*1*). ATP-dependent nucleosome remodelers, which share similar ATPase domains and have an affinity to the nucleosome, are categorized into four subfamilies based on their unique domains and associated subunits: SWI/SNF, ISWI (imitation switch), CHD (chromatin helicase DNA-binding), and INO80/SWR1.

A number of neurodevelopmental disorders have been linked to pathogenic variants in genes encoding chromatin remodelers and associated subunits, including the SWI/SNF subunits ARID1A (MIM: 603024), ARID1B (MIM: 614556), ARID2 (MIM: 609539), SMARCA4 (MIM: 603254), SMARCB1 (MIM: 601607), SMARCE1 (MIM: 603111), SMARCD1 (MIM: 601735), SMARCC2 (MIM: 601734), and DPF2 (MIM: 601671) in Coffin-Siris syndrome (*2-8*); SMARCA2 (MIM: 600014) in Nicolaides-Baraitser syndrome (MIM: 601358) (*9, 10*); BCL11A (MIM: 606557) in Dias-Logan syndrome (MIM: 617101) (*11*); ACTL6B in Intellectual developmental disorder with severe speech and ambulation defects (MIM: 618487) (*12*); ACTB (MIM: 102630) and ACTG1 (MIM: 102560) in Baraitser-Winter syndrome (*13*); CHD subunits CHD4 (MIM: 603277) in Sifrim-Hitz-Weiss syndrome (MIM: 617159) (*14, 15*), CHD7 (MIM:608892) in CHARGE syndrome (MIM: 214800) (*16*), and CHD3 (MIM: 602120) in Snijders Blok-Campeau syndrome (MIM: 618205) (*17*); ISWI subunits BPTF (MIM: 601819) in neurodevelopmental disorder with dysmorphic facies and distal limb anomalies (*18*); and finally, SMARCA1 in Coffin-Siris syndrome-like and Rett syndrome-like phenotypes (*19, 20*).

ISWI is one of the well-studied chromatin remodeling complexes in animal models. Iswi was first identified in *Drosophila melanogaster*, which has a single Iswi ATPase motor protein compared to two in humans (*21-23*). In vertebrates, the ISWI complex is typically composed of 2-4 subunits, including an ATPase, a catalytic subunit, and a scaffolding component involved in target recognition or stabilization. Remarkably, eight different ISWI-containing chromatin remodeling complexes have been identified in humans, including ACF, CHRAC, NoRC, RSF, WICH, SNF2H/NURD/Cohesin, CERF, and NURF (*24-31*). All of these complexes contain either the SNF2L or SNF2H ATPase subunit encoded by *SMARCA1* or *SMARCA5*, respectively. The two proteins share 83% amino acid similarity. Of note, six of the eight complexes use SNF2H exclusively instead of SNF2L, highlighting an indispensable function of SMARCA5. Indeed, morpholino knockdown of the zebrafish *SMARCA5* ortholog results in decreased body size and reduced cardiac ventricle size (*32*). A critical role for SMARCA5 in mammalian development has also been demonstrated in mice where knockout embryos die during preimplantation stages and conditional knockouts lead to significant reductions in body weight, body size, and brain size with cerebellar hypoplasia (*33*). However, a specific syndrome associated with *SMARCA5* rare germline variants has not yet been described. In addition, whereas SMARCA5 has been implicated in neural stem cell proliferation and differentiation (*26, 33, 34*), the spectrum of its functions in the nervous system remains undetermined, with the underlying mechanisms largely underexplored.

Here, we report eleven individuals (six females and six males) from ten unrelated families assembled through international collaborations with germline variants in *SMARCA5*. These individuals share the core presentations of mild developmental delay, postnatal microcephaly, short stature, and mild facial dysmorphia. Using a *Drosophila* model, we found that Iswi loss-of-function (LoF) results in a spectrum of neurodevelopmental abnormalities including reduced body/brain size, dendrite and axon mis-patterning, and behavioral deficits. These findings mirror the key growth and cognitive phenotypes present in affected individuals. More importantly, human wild-type *SMARCA5* but not affected individuals’ variants rescue these phenotypes, confirming their pathogenic role in these models. Together, these data describe a novel syndromic neurodevelopmental disorder caused by pathogenic variants in the *SMARCA5* gene.

## Results

### Clinical presentations of individuals with *SMARCA5* variants

After identifying *de novo* or dominantly inherited variants in *SMARCA5* in each family, we assembled the cohort of twelve individuals through the assistance of GeneMatcher (*35*). Clinical phenotypes overlapped and are summarized in **Table 1**, and detailed data compiled in **Supplemental Table 1**. The majority of the probands (8/10; 80%) had neurodevelopmental disorders with developmental delays in some area, but most delays were mild. Since detailed developmental history information was inadequate for the two adult individuals, their degree of neurodevelopmental delay is unclear. While all probands have normal hearing, mild (N = 4) or severe (N = 2) speech delay was present in six probands (67%; 6/9), with the age of first words ranging from 12 to 27 months. Motor delay was reported in six probands (60%) with the age of walking ranging from 15 to 48 months. Hypotonia was found in five probands (50%) and peripheral hypertonia was noted in individual 4 during infancy. Regarding cognition, four probands had mild intellectual disability and one had severe intellectual disability (50%). A formal diagnosis of autism spectrum disorder was made in only individual 3, and attention deficit disorder/attention-deficit/hyperactivity disorder (ADHD) was detected in three probands (30%). A brain MRI scan was performed for seven probands, and brain CT scan or head ultrasound for two additional probands, showing generally normal morphology, although individual 11 showed mildly delayed myelination and periventricular occipital gliosis and individual 12 showed Chiari malformation type 1. Seizures occurred in two probands (20%). Postnatal microcephaly (head circumference (HC) ≤ −2 SD), short stature (height ≤ −2 SD), and failure to thrive, which were concomitant with feeding difficulties, were present in eight probands (80%), among whom, two individuals had birth weight below −2 SD, one had birth length below −2 SD, and three had birth HC less than −2.5 SD. Two individuals in the cohort, individuals 10 and 11, had normal postnatal height and weight, with individual 11 having a normal head circumference, and individual 10 having macrocephaly. Strikingly, 8 out of 9 probands in this cohort (89%) had some type of abnormalities of extremities, including brachydactyly (2/5), clinodactyly (3/7), 2-3 toe syndactyly (2/7), and hallux valgus/sandal gap (6/7), whereas cardiac, gastrointestinal, and genitourinary defects were uncommon (**Supplemental Table 1**). Individual 2 had multiple cardiac septal defects and a non-compaction dilated cardiomyopathy, with no causative variants in known cardiac genes identified by her clinical exome sequencing. A dermatologic finding, café au lait macule, was identified in two individuals (individuals 3 and 9), who had the recurrent variant, c.1301_1306del, p.(Ile434_Leu435del). Photographs from participants show a number of shared craniofacial features, including blepharophimosis and/or short palpebral fissures with or without almond-shaped eyes (8/10), periorbital fullness (8/9), epicanthal folds (4/10), a wide/high nasal bridge (8/9), a short/flat philtrum (7/9), a thin or tented upper lip (8/10), arched and sparse eyebrows (6/11), medial flaring eyebrows (5/9), and prominent ears (5/10) (**Supplemental Table 1**). Vision abnormalities are reported in 4/12 cases, including myopia, hyperopia, strabismus, and astigmatism. A search the ClinVar database noted one additional individual (ID 599594) with short stature and a *de novo SMARCA5* variant, c.940A>C, p.(Lys314Gln), although no detailed clinical information was available for this subject. Taken together, these data indicated that variants in *SMARCA5* are associated with a variable neurodevelopmental phenotype with predominantly mild developmental delay and intellectual disability, short stature and microcephaly.

**Table 1.** Summary of clinical characteristics associated with heterozygous SMARCA5 variant.

| Clinical findings | # of individuals / assessed | Percentage |
| --- | --- | --- |
| Gender | 5 Female and 6 Male |  |
| **Neurodevelopment** |  |  |
| Prenatal microcephaly | 3/7 | 43% |
| Postnatal microcephaly | 10/12 | 83% |
| Postnatal short stature | 10/12 | 83% |
| Failure to thrive | 8/10 | 80% |
| Hypotonia | 5/10 | 50% |
| Developmental delays | 8/10 | 80% |
| Motor delay | 6/10 | 60% |
| Speech/language delay | 6/9 | 67% |
| Behavioral problems | 3/10 | 30% |
| **Facial feature** |  |  |
| Blepharophimosis/short palpebral fissures | 8/10 | 80% |
| Epicanthal folds | 4/10 | 40% |
| Periorbital fullness | 8/9 | 89% |
| Wide/high nasal bridge | 8/9 | 89% |
| Short/flat philtrum | 7/9 | 78% |
| Thin or tented upper lip | 8/10 | 80% |
| Medial flaring of the eyebrows | 5/9 | 56% |
| High arched and sparse eyebrows | 6/11 | 50% |
| Prominent ear | 5/9 | 55% |
| **Other abnormalities** |  |  |
| Limb abnormalities | 8/9 | 89% |
| Eye/Vision | 4/12 | 33% |
| Seizures | 2/10 | 20% |

### Identification of *SMARCA5* variants

Exome sequencing of the ten families revealed nine different heterozygous novel variants in *SMARCA5* (NM_003601.3), including eight variants confirmed to be *de novo* in nine individuals. Notably two unrelated individuals were found to carry the same *de novo* variant, c.1301_1306del, p.(Ile434_Leu435del). One missense variant, c.1711G>C, p.(Ala571Pro), was identified in a three-generation pedigree. These nine variants include one splice-altering variant, c.802-2A>G; one in-frame deletion, c.1301_1306del, p.(Ile434_Leu435del); and seven missense variants, c.1130A>T, p.(Asp377Val); c.1682C>A, p.(Ala561Asp); c.1711G>C, p.(Ala571Pro); c.1775G>A, p.(Arg592Gln); c.1855G>C, p.(Glu619Gln); c.2207G>A, p.(Arg736Gln); and c.2677G>A, p.(Glu893Lys) (**Figure 1A**). All variants were absent in population genomics resources (i.e., 1000 Genomes Project, ESP6500SI, and gnomAD) and our in-house dataset of >10,000 exomes. All seven missense variants were predicted to be deleterious by multiple bioinformatic prediction algorithms (SIFT, LRT, MutationTaster, CADD, etc.) (**Table S2**). RT-PCR followed by Sanger sequencing of cDNA from fibroblasts and a lymphoblastoid cell line (LCL) obtained from individual 1 with the splice-altering variant demonstrated that it causes exon 7 skipping, leading to an in-frame deletion of 52 amino acids (p.Ala268_Lys319del) in the highly conserved ATPase domain (**Figure 1B**). These missense variants and the small deletions were mapped to a protein schematic and eight variants appear to cluster within or around the helicase domains, whereas p.(Arg736Gln) and p.(Glu893Lys) are located in the HAND-SANT-SLIDE (HSS) domains (**Figure 1A**). Of note, all the variants reside in protein locations intolerant to change as calculated by the MetaDome server (**Figure 1A and Table S2**).

**Figure 1.**
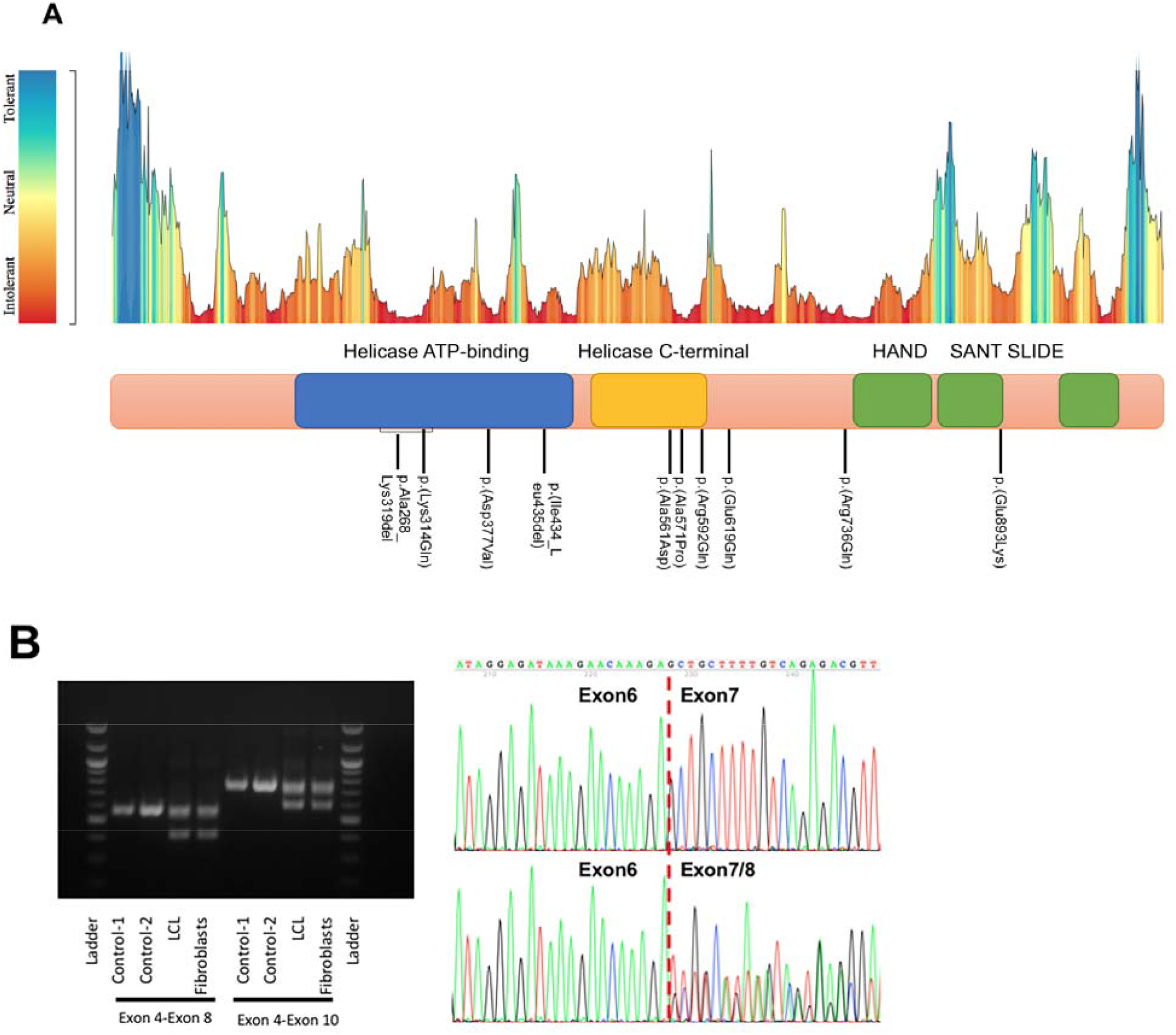
Molecular genetic findings. **(A)** An intolerance landscape plot generated by MetaDome for *SMARCA5* variant analysis (upper panel) and a schematic outline of SMARCA5 protein showing conserved variants identified in individuals 1-12 and ClinVar at the bottom. (**B**) RT-PCR analysis of individual 1’s fibroblasts and lymphoblastoid cell line showing the *SMARCA5* exon 7 skipping in both cell types due to a splice-altering variant.

To explore the structural impact of the six missense variants within or around the helicase domains, the recently published cryo-electron microscopy three-dimensional structure (*36*) of wild-type (WT) SMARCA5 incorporated with the nucleosome complex was obtained (PDB: 6ne3). SMARCA5 binds to DNA in the nucleosome through its ATPase helicase domains, which form a positively charged cleft on the binding surface (**Figure S1A**). The p.(Lys314Gln) and p.(Asp377Val) changes result in loss of the positive charge, and the p.(Ala561Asp) change introduce a negatively charged side chain, which may diminish its anchor of DNA binding (**Table S2; Figure S1B-C**). Similarly, the p.(Ala571Pro), p.(Arg592Gln), and p.(Glu619Gln) changes may disturb hydrogen bonds and structural stability, therefore possibly affecting interaction with DNA, histone, or ADP ligand binding (**Table S2; Figure S1B-C**). Recent studies have demonstrated the acidic patch, a negatively charged structure on the surface of the nucleosome, is critically important for recruiting chromatin remodelers, and an acidic patch binding (APB) motif in SMARCA5 has been discovered (*37*). Notably, the substitution of Arg736, one of the five residues in the APB motif, with alanine in SMARCA5 disrupted its nucleosome sliding activity (*38*). It seems reasonable to speculate that the replacement of Arg736 with glutamine, similar loss of positive charge and the side chain changing from a large residue to a small one, could similarly alter its remodeler activity. The three-dimensional structure of the HSS domains of WT human SMARCA5 was obtained by using homology modeling with SWISS-MODEL. The X-ray structure of *Drosophila* Iswi at a 1.9 AL resolution was used as a template as it demonstrates 81.25% identity with human SMARCA5 (PDB: 1ofc) (*39*). As shown on **Figure S1D**, residue Glu893 was mapped onto the SANT domain helix 3 (SANT3), a recognition helix which makes contacts with unmodified histone tails and has an overall negatively charged surface. A change from a negatively charged side chain to a positively charged one [p.(Glu893Lys)] in the SANT motif, which is predicted to disrupt a local salt bridge, may affect SANT motif substrate recognition ability to contact histone, and in turn to hinder ATPase binding.

### *SMARCA5*’s ortholog *Iswi* plays a key role in *Drosophila* larval development

To gain independent support for pathogenicity of the variants identified in humans and investigate whether SMARCA5 is required for normal development, we studied the *Drosophila* gene *Iswi (Imitation SWI*), which is orthologous to human *SMARCA1* or *SMARCA5*. We first assessed the functional effects of an *Iswi* loss-of-function (LoF) mutant, *Iswi*^*2*^, on larvae development (*40*) and found that *Iswi*^*2*^ homozygous larvae died before the end of larval stage. For those that survived to 3^rd^ instar, body size was substantially smaller than WT larvae (**Figure 2A-B**). It was previously reported that Iswi is important for chromosome decondensation during mitosis, and *Iswi* LoF leads to mitotic recombination defects (*40, 41*). To investigate whether Iswi affects cell mitosis, we performed immunostaining with phosphorylated Histone H3 (pH3), a proliferation maker for mitotic cells. We observed that in the brain lobes, the number of pH3^+^ positive cells was significantly decreased in *Iswi*^*2*^ larvae (**Figure 2C-D**). We next asked whether Iswi may play a role in neuronal development. To accomplish this, we used *Drosophila* dendritic arborization (da) sensory neurons, which provide a suitable system to study neuronal development and dendritic morphology. There are four classes of da neurons, with class IV da (C4da) neurons extending the most complex and highly branched dendrites (*42*). We found that in *Iswi*^*2*^, the dendrite complexity of C4da neurons was decreased. Both the total dendrite length and dendritic branch number exhibited a significant reduction (**Figure S2B-D**), indicating that Iswi is important for dendrite morphogenesis. Furthermore, *Iswi* LoF also caused a tiling defect between C4da neurons ddaC and v’ada. Tiling refers to a phenomenon in which the dendrites of adjacent neurons establish complete, but non-redundant interaction to occupy the space between the cell bodies, as observed in WT larvae (*43*). In *Iswi*^*2*^ larvae, however, the space is not fully covered, as there is a significantly enlarged gap in the dendritic field between ddaC and v’ada (**Figure 2E-F**). These results suggest that Iswi plays a pleiotropic role during fly larval development, highlighting its involvement in neural proliferation and morphogenesis.

**Figure 2.**
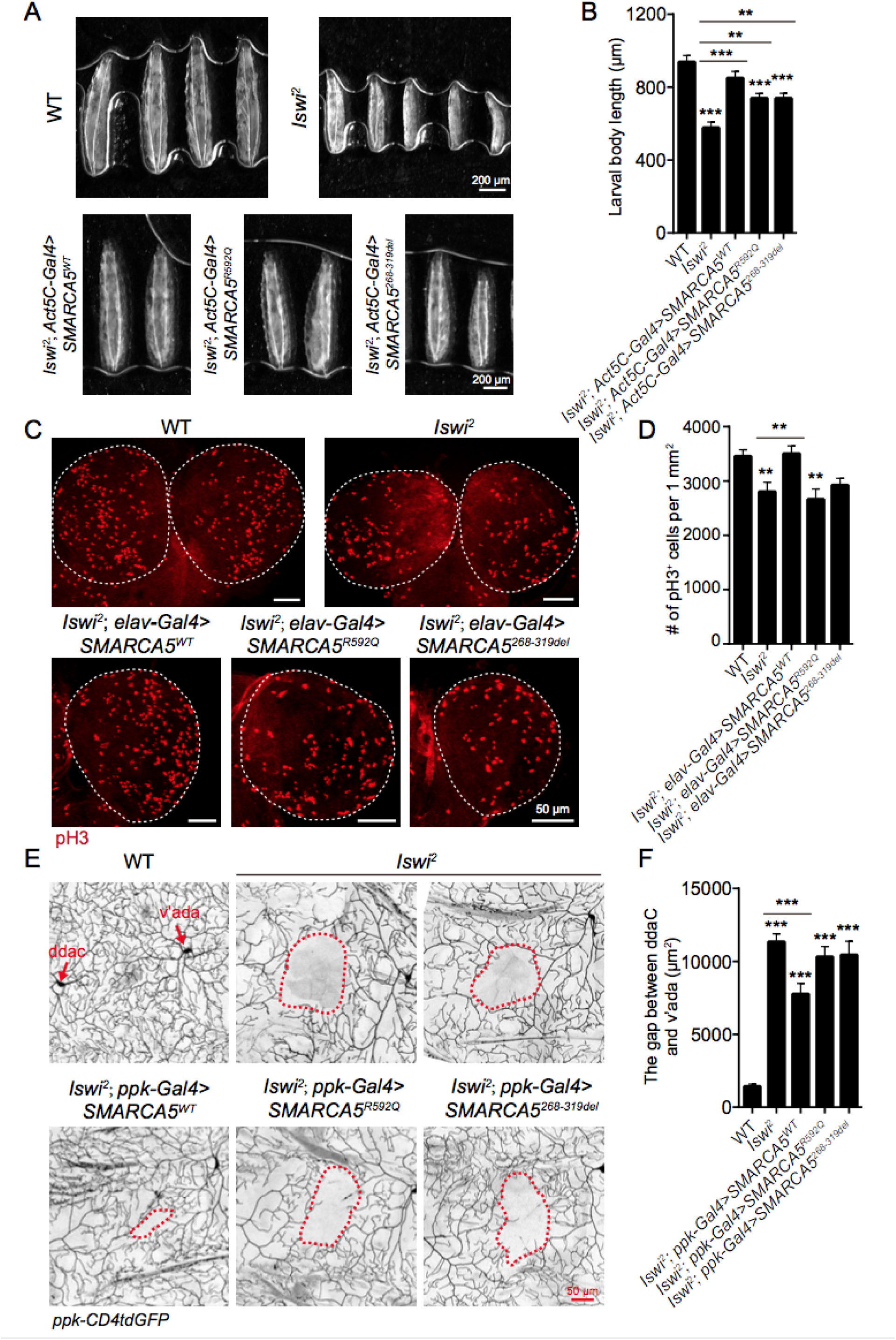
*Iswi* loss of function leads to developmental defects in *Drosophila* larvae. **(A-B)** Compared with WT, larval body size is significantly smaller in *Iswi*^*2*^ 3^rd^ instar larvae, which can be rescued by the expression of SMARCA5^WT^ by *Act5C-Gal4*. (A) Images showing larvae of different genotypes. Scale bar: 200 μm. (B) Quantification of larval body length. N = 30 to 41 larvae. **(C-D)** In the brain lobes of *Iswi*^*2*^ larvae, the number of pH3^+^ positive mitotic cells is significantly decreased. N *=* 12 to 17 brain lobes. Expressing SMARCA5^WT^ by *elav>Gal4* is sufficient to rescue the decreased proliferation in *Iswi* LoF larvae. (C) The brains were dissected from 3^rd^ instar larvae and stained with pH3. Scale bar: 50 μm. (D) Quantification of pH3^+^ positive mitotic cells, *P* = 0.0090. **(E-F)** In *Iswi*^*2*^, C4da ddaC and v’ada neurons exhibit dendritic tiling defects. SMARCA5^WT^, but not patient variants, partially rescues the tiling defects. Scale bar: 50 μm. (E) The red dashed circles outline the gap area between C4da ddaC and v’ada neurons. (F) Quantification of the gap in C4da neuron dendritic field, N = 17 to 34 dendritic fields from 4 to 5 larvae. All data are mean ± SEM. The data were analyzed by one-way ANOVA followed by Dunnett’s multiple comparisons test, \*\**P* < 0.01, \*\*\**P* < 0.001.

We next expressed *Iswi* RNAi under the control of *elav-Gal4*, a pan-neuronal driver (*44*), to determine whether Iswi is cell-autonomously required in neurons. Indeed, neural-specific knockdown of *Iswi* leads to significantly smaller brain size in adult flies (**Figure 3A-B**). Moreover, in the *Iswi* knockdown brains, we observed severe structural deficits of the mushroom body, the learning and memory center of the fly (*45*). Compared to control, the volume of both the vertical lobe and the horizontal lobe is drastically reduced after *Iswi* knockdown, with bilaterally missing α/β lobes and reduction of the γ lobe (**Figure 3C-E**). Consistent with these findings, Gong *et al*. reported that *Iswi* knockdown in flies results in mushroom body morphologic abnormalities, as well as disrupted sleep, circadian rhythmicity, memory, and social behaviors, suggesting that Iswi is required during development for numerous adult behaviors (*46*).

**Figure 3.**
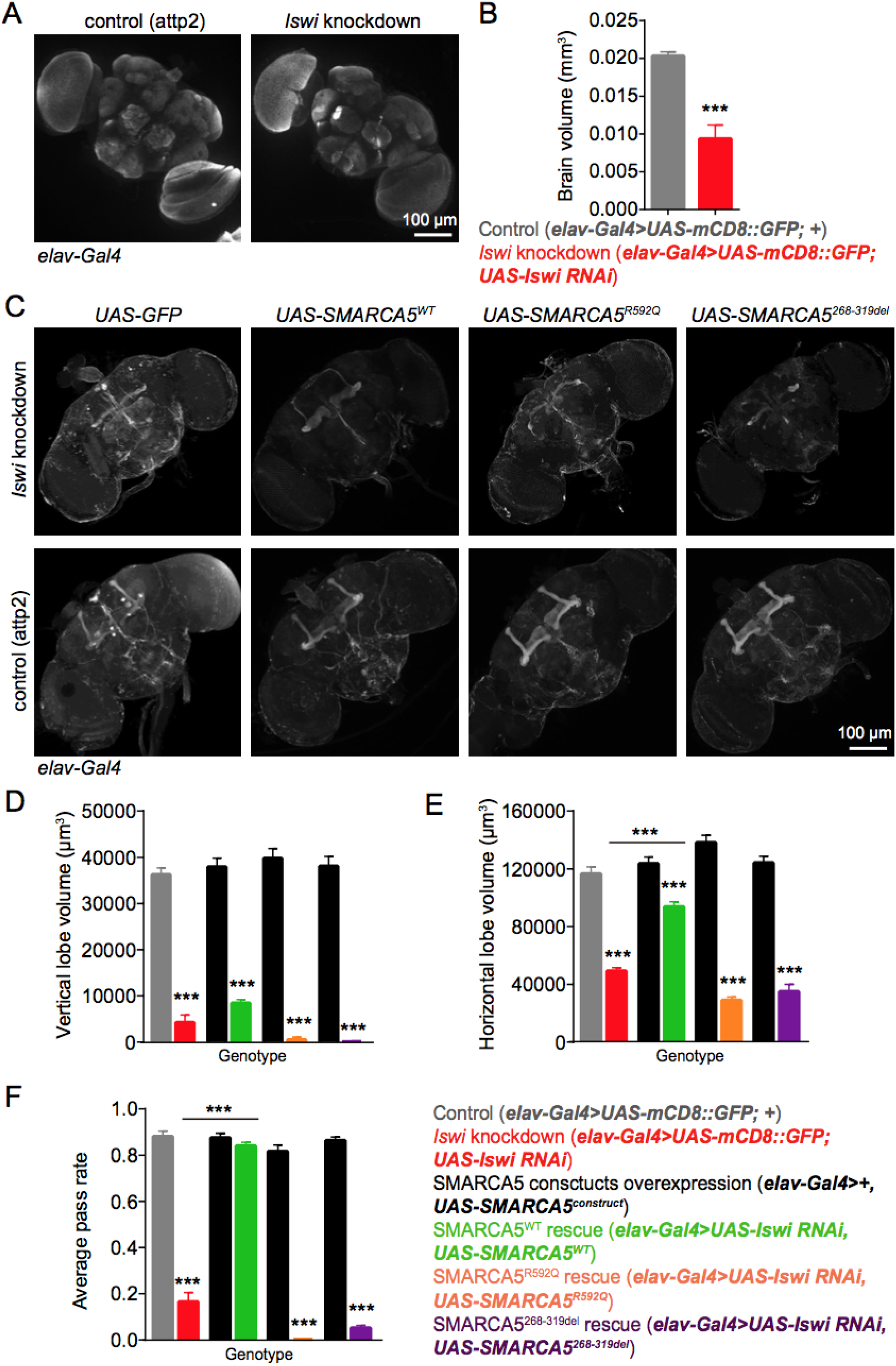
*Iswi* neural-specific knockdown causes abnormal brain structure and motor deficits in adult files. **(A-B)** *Iswi* neural-specific knockdown leads to smaller brain size compared with control flies. (A) Representative images of fly brains of control and *Iswi* knockdown flies. Scale bar: 100 μm. (B) Quantification of adult fly brain volume. *N* = 6, *P* = 0.0002. **(C-E)** *Iswi* neural-specific knockdown affects mushroom body structure in adult flies. While expressing SMARCA5^WT^ partially rescues the structure, the patient variants are not able to rescue mushroom body morphology. N = 11 to 35 brains. (C) Representative images of fly brains from different groups. Mushroom bodies are marked by FasII staining. Scale bar: 100 μm. (D) Quantification of vertical lobe volume. (E) Quantification of horizontal lobe volume. SMARCA5^WT^ rescue significantly increases the horizontal lobe volume in *Iswi* knockdown flies. **(F)** The negative geotaxis test shows that *Iswi* knockdown leads to locomotion deficit in flies. Expression of SMARCA5^WT^ in *Iswi* knockdown flies is able to rescue the climbing deficit to control levels, while SMARCA5^R592Q^ and SMARCA5^268-319del^ mutants fail to rescue. N = 3 to 12 groups, with each group contain 10 flies. All data are mean ± SEM. The data were analyzed by unpaired t-test or one-way ANOVA followed by Tukey’s multiple comparison test, \*\**P* < 0.01, \*\*\**P* < 0.001.

As motor delay was reported in several patients, we next examined the locomotor function in *Iswi* knockdown flies by the negative geotaxis assay (a climbing test). We found that while ∼80% of control flies climbed to or past the target distance, only a small portion of *Iswi* knockdown flies (less than 20%) showed normal climbing ability (**Figure 3F**), indicating that neural knockdown of *Iswi* severely impaired the motor skills in flies. Taken together, these data indicate that Iswi is necessary for brain development and function in flies.

### SMARCA5 WT but not mutants rescue the defects observed after *Iswi* LoF

To determine the functional consequences of the *SMARCA5* variants identified in individuals 1 and 2, we generated transgenic flies with inducible expression of WT SMARCA5 (SMARCA5^WT^), SMARCA5^R592Q^ and SMARCA5^268-319del^, respectively. We found that these variants did not change SMARCA5’s localization in the nucleus (**Figure S2A**). We investigated whether human SMARCA5^WT^ and variants could rescue the defects observed in *Iswi*^*2*^ larvae and neural-specific *Iswi* knockdown flies. First, we expressed SMARCA5^WT^ and the two human variants in *Iswi*^*2*^ larvae under the control of *Act5C-Gal4*, which drives the expression ubiquitously (*47*), and found that SMARCA5^WT^ expression was sufficient to rescue the larval body size similar to WT. In contrast, SMARCA5^R592Q^ and SMARCA5^268-319del^ only partially rescued the body length in *Iswi*^*2*^ larvae (**Figure 2A-B**). Second, we examined whether the expression of WT or mutant SMARCA5 rescued the decreased cellular proliferation observed in *Iswi*^*2*^ larval brain lobes. When using the *elav-Gal4* driver to express SMARCA5^WT^ in *Iswi* LoF larvae, we found that SMARCA5^WT^ robustly rescued the proliferation deficits, while no rescue was observed when expressing the two patient variants.

Third, we expressed SMARCA5^WT^ in C4da neurons of *Iswi*^*2*^ larvae using the C4da neuron specific *ppk-Gal4* driver (*42*), and found that SMARCA5^WT^ substantially rescued the tiling deficit between ddaC and v’ada, resulting in significantly decreased gap area in the dendritic field. Perhaps because *Drosophila Iswi* is the ortholog of the human *SMARCA1* and *SMARCA5*, expressing SMARCA5 only partially restored the neurodevelopmental defects in *Iswi*^*2*^ (**Figure 2E-F**). Notably, both human *SMARCA5* variants completely failed to rescue the tiling phenotype in *Iswi*^*2*^ (**Figure 2E-F**). Interestingly, overexpressing SMARCA5^WT^ in WT larvae reduced dendritic complexity (**Figure S2B-D**), similar to the phenotype observed in *Iswi*^*2*^, suggesting that the precise control of Iswi expression is crucial for normal dendrite morphogenesis. While the expression of SMARCA5^R592Q^ also caused impaired dendrite outgrowth and branching, the dendrite length and branch number in SMARCA5^268-319del^ overexpressing neurons were comparable to WT flies without transgenic expression of human SMARCA5 (**Figure S2B-D**). We did not observe dendritic tiling defects in any of the three overexpression transgenic lines. These results suggest that although both variants jeopardized normal function of SMARCA5, they appear to differ in their mutagenic strength, and may not be complete LoF.

Finally, human WT SMARCA5 partially reversed the mushroom body morphologic abnormalities and fully rescued locomotion defects induced by neural-specific *Iswi* knockdown. We found that the expression of SMARCA5^WT^ partially restored horizontal lobe volume of the mushroom body, although it had little effect on the vertical lobes (**Figure 3C-E**). Comparatively, expressing either SMARCA5^R592Q^ or SMARCA5^268-319del^ was unable to rescue the morphological defects. Remarkably, in the negative geotaxis test, SMARCA5^WT^ fully rescued the climbing deficit in *Iswi* knockdown flies, resulting in similar motor performance as control flies, while the patient variants failed to improve climbing capacity (**Figure 3F**).

## Discussion

We describe a novel syndromic neurodevelopmental disorder caused by *de novo* or dominantly inherited variants in *SMARCA5*, a gene that encodes an ATPase motor protein in the ISWI chromatin remodeler complex and which is extremely intolerant to both LoF (pLI = 1) and missense (Z = 5.42) variants based on the gnomAD population database. We identified six missense variants, one splice-altering variant that lead to exon skipping and in-frame deletion, and one recurrent in-frame deletion in eleven individuals from nine unrelated families from around the world. The individuals in our study demonstrated a broad range of clinical features, as is not uncommon in neurodevelopmental disorders associated with pathogenic variants in aforementioned chromatin remodeling complexes (i.e., SWI/SNF, CHD, and ISWI) that vary from isolated autism to syndromic intellectual disability (*5, 17, 48, 49*). The most consistent clinical findings in our cohort are postnatal microcephaly and short stature observed in 10/12 individuals, and developmental delay in 8/10 individuals, although again, the severity of these delays varies widely from mild to severe (individual 7). Interestingly, individual 10 has macrocephaly with a variant outside the helicase C-terminal domain and individual 11 is normocephalic with a variant in HSS domains; both of these individuals have delays in motor and speech development but normal stature, illustrating phenotypic variability. Conversely,individuals 1 and 8 have postnatal microcephaly and short stature but normal development with variants in the helicase ATP-binding domain. Varying degrees of developmental delay have been consistently reported in neurodevelopmental disorders associated with pathogenic variants involved in chromatin remodeling complexes. Similarly, varying head size ranging from microcephalic to macrocephalic is also frequently observed, for example, in *SMARCB1* and *CHD3* (*49, 50*). It is widely acknowledged that different variants within different functional domains of a gene can cause different phenotypes. Although the numbers are relatively small, when comparing phenotypes to identified variants, no clear genotype-phenotype correlation was established in this cohort. It is noteworthy that variants in *SMARCA1*, which is highly homologous to *SMARCA5*, have been associated with distinct phenotypes such as Rett syndrome, Coffin-Siris-like syndrome, and sporadic schizophrenia (*19, 20, 51*). This can be explained by mutually exclusive presence of SMARCA1 and SMARCA5 in the different ISWI complexes that often have overlapping as well as complex-specific functions.

*SMARCA5* is located at 4q31.21, a region overlapping with the deletions observed in the 4q deletion syndrome, which is characterized by a broad clinical spectrum and phenotypic variability (*52*). Review of the literature revealed that two subjects have interstitial deletions encompassing the *SMARCA5* gene. One patient with 14 Mb deletion (4q28.3-31.23) has development delay, growth failure, ventricular septum defect, and craniofacial dysmorphisms (*53*); whereas the other subject with a 6 Mb deletion has only mild speech delay (*54*). Searching the Decipher database returned fourteen participants that have deletions containing *SMARCA5* with a range of heterogeneity in clinical findings. Since all of these deletions are larger than 6 Mb and include many other genes, it is unclear how haploinsufficiency of *SMARCA5* contributes to their specific phenotypes, but it suggests that heterozygous *SMARCA5* deletion is compatible with life. Based on our findings, we propose that *SMARCA5* should be considered in the context of 4q deletion syndrome.

In general, this novel *SMARCA5*-associated syndrome reported here appears to result in a less severe phenotype when compared to other neurodevelopmental disorders due to pathogenic variants in *ARID, SMARC*, and *CHD* gene families, where almost all of the variants occur *de novo* and parent-child transmission is rarely documented (*49, 55*). We present here a three-generation pedigree (for individuals 4-6) segregating a deleterious missense variant. It is interesting to note that individual 4’s maternal grandfather also has short stature (indicated in the pedigree; **Figure S3**); however, he has a pathogenic variant in *TLK2* (NM_006852.3:c.364C>T, p.(Arg122*)) which is associated with “Mental retardation, autosomal dominant 57” (OMIM **#** 618050) which also includes short stature. This variant is not inherited by individuals 5 and 6 as confirmed by exome sequencing. Although *SMARCA5*-associated neurological complications are generally mild, postnatal proportionate short stature and microcephaly are rather predominant with standard deviations ranging from −2 to −4.78 for height, and −2 to −6.21 for head circumference, in 8/10 probands. Individuals presented here have craniofacial dysmorphisms and some facial features overlap with *SMARCA2*-associated blepharophimosis intellectual disability syndrome (*56*), including blepharophimosis, short palpebral fissures, epicanthal folds, high arched and sparse eyebrows, and short metacarpals. Overall, individuals with the *SMARCA5*-associated phenotypes appear to share similar dysmorphic features and clinical histories, including blepharophimosis, short palpebral fissures, periorbital fullness, wide/high nasal bridge, a thin or tented upper lip, short/flat philtrum, high arched, sparse, or medial flaring eyebrows, and hallux valgus, but each of the shared clinical findings is relatively non-specific. Additional individuals and further assessment will be required to further clarify the clinical spectrum and constellation of features associated with this novel disorder to subsequently assist in the clinical diagnosis; however, *SMARCA5* gene sequencing should be considered in patients with failure to thrive, short stature, microcephaly, and dysmorphic features.

Our phenotypic analyses in the fly *Iswi* LoF models and human patients emphasize the pleiotropic and critical role of SMARCA5 and the SNF2H-containing chromatin remodeling complexes in nervous system development. We observed that LoF mutants of its *Drosophila* ortholog *Iswi* led to decreased larval body size and sensory dendrite complexity, together with a tiling defect. Apart from its previously reported function in cell proliferation/stem cell self-renewal (*57*) and hence smaller head/microcephaly, we identified its requirement in post-mitotic neurons. Specifically, we found that *Iswi* LoF leads to defects in sensory neuron dendrite tiling and mushroom body development, highlighting its underappreciated role in neural circuit assembly (*46*). This likely contributes to the behavioral deficits such as impaired locomotion in the *Iswi* knockdown flies, and intellectual disability, hypotonia and speech delay in patients. Nevertheless, the patient variants, SMARCA5^R592Q^ and SMARCA5^268-319del^, largely failed to replace Iswi in flies, highlighting the functional effect of the variants that we identified. Given that both variants could partially rescue the reduced body size in *Iswi*^*2*^, and that the overexpression of SMARCA5^R592Q^ in neurons affects their dendrite morphology, these two variants may not be complete LoF alleles and may retain partial function allowing it to interact with the dendrite morphogenesis machinery. Modeling variants in the three-dimensional structure suggests that they may disrupt interactions of SMARCA5 with the nucleosome and in turn affect ATPase binding. An emerging question is how SMARCA5 and more generally the chromatin remodelers exert their diverse functions under various developmental scenarios. Therefore, an imminent task is to identify the DNA substrates processed by the remodelers at a particular timepoint and in a specialized tissue niche, which could be potential therapeutic targets. Altogether, our functional studies of the *SMARCA5* missense variants suggest that they are hypomorphic alleles. This could potentially explain the similar but generally mild phenotypes in the affected individuals.

In summary, we identify and phenotypically characterize a novel neurodevelopmental syndrome that overlaps with conditions caused by variants in different subunits of chromatin remodeling complexes. Frequent features include postnatal short stature and microcephaly, in addition to shared dysmorphia. Our findings in a fly *in vivo* model highlight the important and previously underappreciated role of the ISWI family proteins in dendrite morphogenesis, neural circuit formation, and diverse behaviors. In conclusion, this study expands the spectrum of ISWI-related disorders and extends the tally of causative genes in neurodevelopmental disorders.

## Data Availability

Patient variants will be submitted to a public database. All patient materials may be obtained through an MTA.

---

**Figure S1.**
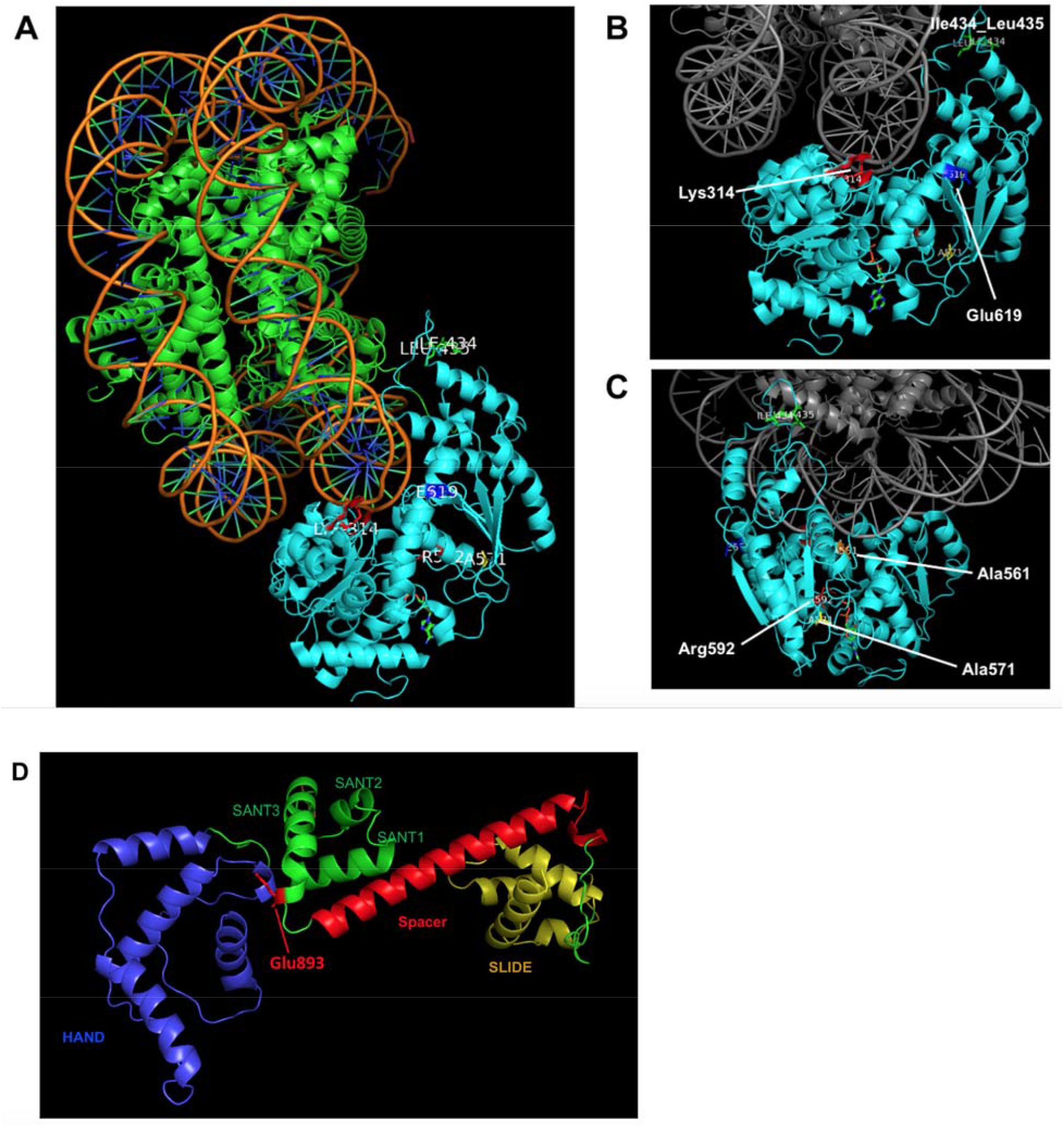
Three-dimensional structure analysis. **(A)** SMARCA5 ATPase domain (cyan) is shown to interact with DNA double helix and histones (PDB: 6ne3). (**B-C**) *SMARCA5* variants mapped on the ATPase domain (cyan) with DNA and histones (grey). (**D**) Three-dimensional structure modeling using homology modeling with SWISS-MODEL and SMARCA5 HSS domain variant Glu893 mapped on a *Drosophila* Iswi protein structure (PDB: 1ofc). Different colors denote different domains.

**Figure S2.**
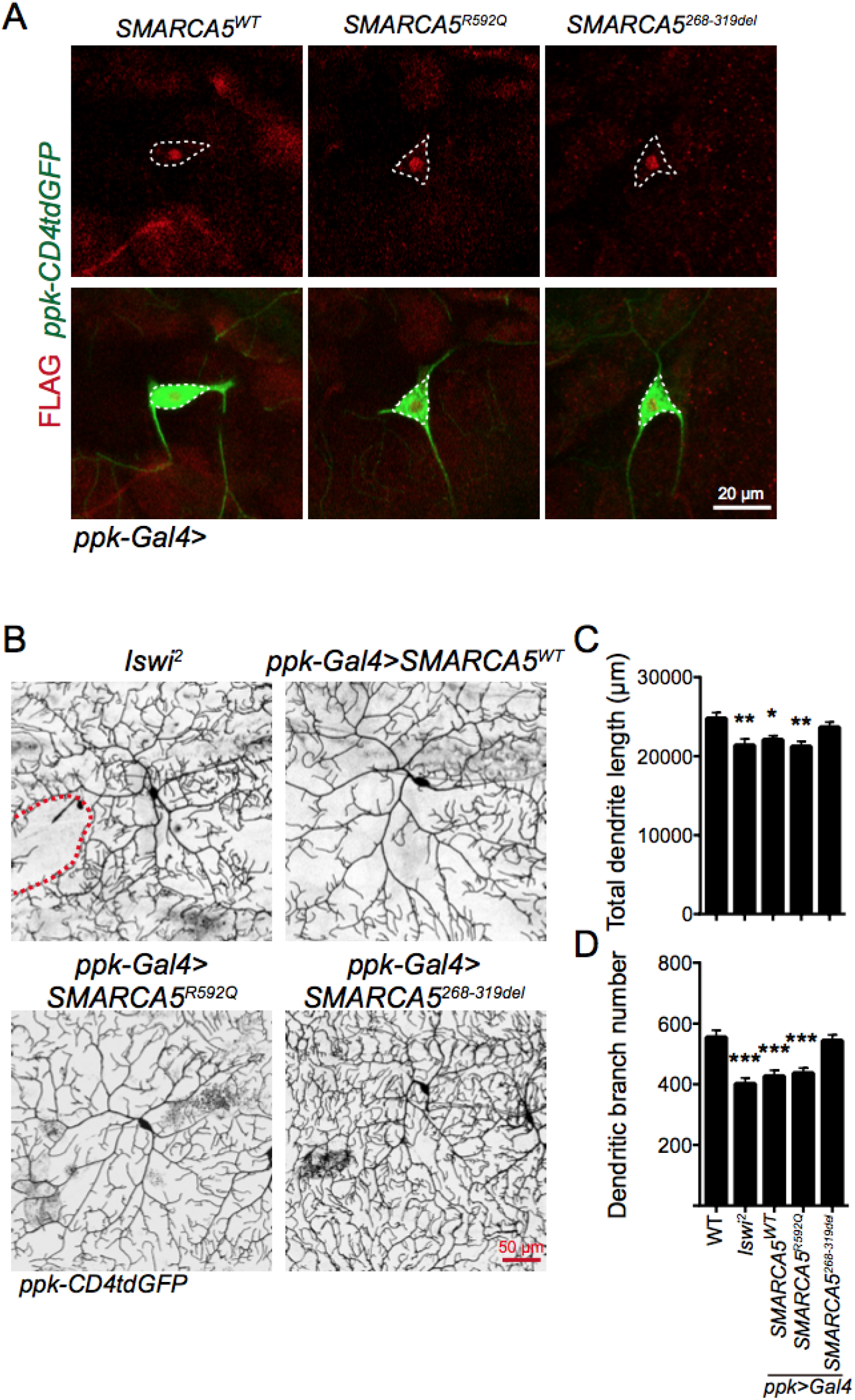
SMARCA5 localization and their overexpression effect on dendrite branching. **(A)** The body walls from SMARCA5^WT^, SMARCA5^R592Q^ and SMARCA5^268-319del^ expressing larvae were dissected and stained for FLAG. Confocal images showed that in C4da neurons, both the WT and human mutant SMARCA5 are mainly localized in the nucleus. Scale bar: 20 μm. **(B-D)** In 3^rd^ instar larvae, *Iswi* LOF reduces dendrite outgrowth and branching in C4da neurons. Their dendrite complexity also exhibits a significant decrease in SMARCA5^WT^ and SMARCA5^R592Q^ but not SMARCA5^268-319del^ overexpressing C4da neurons. N = 18 to 22 neurons from 4 to 6 larvae. (B) Images showing C4da neurons from different transgenic flies. The red dashed line marks the gap area between C4da ddaC and v’ada neurons in *Iswi*^*2*^. Scale bar: 50 μm. (C) Quantification of total dendrite length. (D) Quantification of dendritic branch number. All data are mean ± SEM. The data were analyzed by one-way ANOVA followed by Dunnett’s multiple comparisons test, \**P* < 0.05, \*\*\**P* < 0.001.

**Figure S3.**
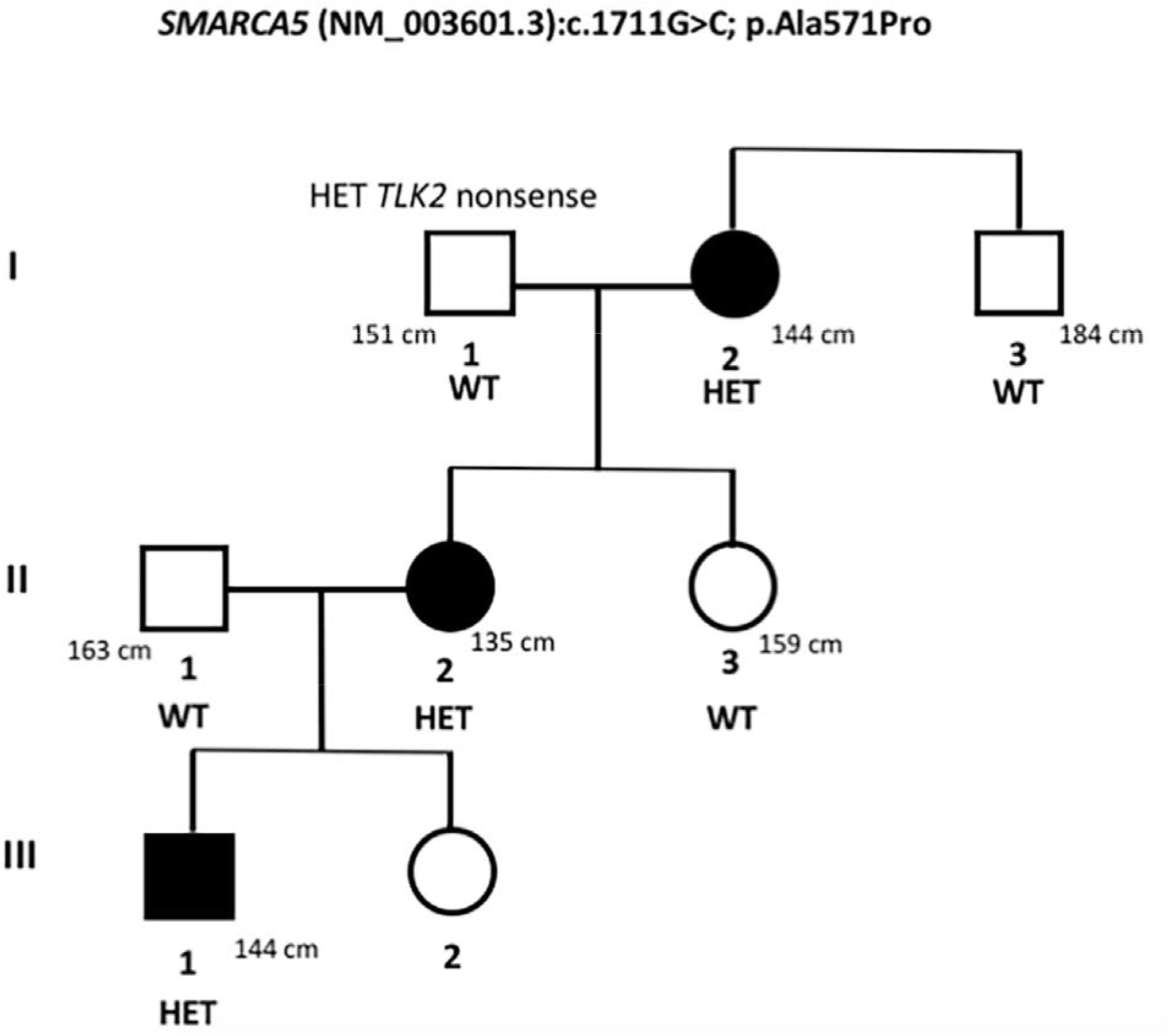
Three-generation family pedigree.

## Methods and Materials

### Research participants

Informed consent was obtained from all the families according to protocols approved by local institutional review boards and human research ethics committees. Permission for clinical photographs was given separately.

### Genetic analysis

Genomic DNA was extracted from whole blood from the affected children and their parents. Exome sequencing was performed with a variety of standard capture kits and data analysis was performed independently. In patients 9 and 11, exome sequencing was performed in the framework of the German project “TRANSLATE NAMSE”, an initiative from the National Action League for People with Rare Diseases (Nationales Aktionsbündnis für Menschen mit Seltenen Erkrankungen, NAMSE) facilitating innovative genetic diagnostics for individuals with suggested rare diseases. The coding sequence of *SMARCA5* in individual 1 was amplified by RT-PCR using primers 5’-TTGCACTCGATTTGAAGACTCT-3’ and 5’-AGTTAAGAAGTGACCACAGCTC-3’.

### Fly stocks

*Ppk-CD4tdGFP, ppk-Gal4, UAS-Iswi RNAi, elav-Gal4, UAS-mCD8::GFP* and *P*{*CaryP*}*attP2* have been previously described (*46, 58*). To generate the *UAS-SMARCA5*^*WT*^, *UAS-SMARCA5R*^*592Q*^ and *UAS-SMARCA5*^*268-319del*^ stocks, the coding sequences were cloned into a pACU2 vector, and the constructs were then injected (Rainbow Transgenic Flies, Inc).

### Live imaging in flies

Live imaging was performed as described (*59*). In brief, embryos were collected for 2-24 hours on yeasted grape juice agar plates and were aged at 25 °C. At 96 h after egg laying, larvae were mounted in 90% glycerol under coverslips sealed with grease and imaged using a Zeiss LSM 880 microscope. Neurons were reconstructed with Neurostudio for dendrite morphology analyses.

### Immunohistochemistry

3^rd^ instar larvae were dissected, and tissues were subjected to immunostaining as described according to standard protocols. The following antibodies were used: mouse anti-FLAG antibody (F3156, 1:1000, Sigma), rat anti-Histone H3 (phospho S28) antibody (ab10543, abcam, 1:500), and fluorescence-conjugated secondary antibodies (1:1000, Jackson ImmunoResearch).

### Negative geotaxis test

Negative geotaxis was performed as described previously (*60*). Three days after eclosion, 10 flies of the same genotype were placed in a 20-cm high vial. After 2-min adaptation, the vial was gently tapped against the table so the flies would be knocked to the bottom of the vial. The pass rate is defined as the percentage of flies climbing beyond the 8 cm mark within 10 seconds. Each vial was assayed 10 times and the average pass rate is defined as the average across 10 trials. Male and female flies were picked randomly.

## Acknowledgements

We thank all of the families involved in this study for their participation. Research reported in this publication was supported in part by the Roberts Collaborative Functional Genomics Rapid Grant (to D.L. and M.A.D.) from CHOP, and Institutional Development Funds (to H.H.) from CHOP. This work was supported by NIH grants DP2 NS111996 (to M.S.K.) and T32 HL07953 (to N.N.G), and funding from the Burroughs Wellcome Fund (to M.S.K). We are grateful for funding support for the Undiagnosed Diseases Program Victoria from the Harbig Family Foundation and the Murdoch Children’s Research Institute. The research conducted at the Murdoch Children’s Research Institute was supported by the Victorian Government’s Operational Infrastructure Support Program. Sequencing and analysis of individual 2 were provided by the Broad Institute of MIT and Harvard Center for Mendelian Genomics (Broad CMG) and was funded by the National Human Genome Research Institute, the National Eye Institute, and the National Heart, Lung and Blood Institute grant UM1 HG008900 and in part by National Human Genome Research Institute grant R01 HG009141.

## Ethics Declaration

All patients’ families from the different institutions agreed to participate in this study and signed appropriate consent forms, including the Children’s Hospital of Philadelphia, Murdoch Children’s Research Institute, Great Ormond Street Hospital, Tel Aviv University, McMaster University, Nationwide Children’s Hospital, Technical University Munich, University of California, Los Angeles, and Phoenix Children’s Hospital. The Institutional Review Board of the Children’s Hospital of Philadelphia approved this study.

